# Acute myocardial infarction releases more troponin per unit of late gadolinium enhancement mass compared to acute myocarditis

**DOI:** 10.64898/2026.02.16.26346430

**Authors:** Manoj Rajamohan, Ashleigh Dind, Martin Ugander, Gemma Figtree, Rebecca Kozor

## Abstract

**Background:** Both acute myocardial infarction (AMI) and acute myocarditis are characterised by cardiac troponin release as a marker of cardiomyocyte injury. While peak troponin is widely accepted as a surrogate marker for infarct size in AMI, its relationship with myocardial injury in acute myocarditis is unclear. This study aimed to quantify and compare the association between peak high-sensitivity cardiac troponin and cardiovascular magnetic resonance (CMR) late gadolinium enhancement (LGE) extent in patients with AMI versus acute myocarditis.

**Methods:** Patients undergoing CMR imaging and measurement of high-sensitivity cardiac troponin I during hospital admission were retrospectively included. LGE extent was quantified in grams using the semi-automated expectation-maximization weighted intensity algorithm (EWA).

**Results:** Compared to patients with acute myocarditis (n=47), patients with AMI (n=49) had higher peak troponin levels (median [interquartile range] 32,470 [3,109–104,699] vs 7,295 [1,857–22,550] ng/L, p=0.002), larger LGE extent (25 [13–56] vs 10 [6–17] g, p<0.001), and lower left ventricular ejection fraction (45 [36– 52] vs 55 [49–58] %, p<0.001). Peak troponin was moderately positively correlated with LGE extent in both AMI (rho=0.56, p<0.001) and acute myocarditis (rho=0.58, p<0.001). However, the ratio of peak troponin to LGE mass was higher in AMI compared to acute myocarditis (1,299 [419–3233] vs 909 [310–1446] ng/L/g, p=0.02).

**Conclusions:** Peak cardiac troponin correlates positively with LGE extent in both AMI and acute myocarditis, but the magnitude of LGE and LV systolic dysfunction is greater in AMI. Also, AMI typically has an approximately 40% greater amount of troponin release per unit LGE mass compared to acute myocarditis. This suggest that troponin-based estimates of myocardial injury size estimated by LGE are not directly interchangeable between ischaemic and inflammatory myocardial diseases.

## INTRODUCTION

Acute myocardial infarction (AMI) and acute myocarditis are potentially life-threatening cardiovascular conditions characterised by cardiac troponin release into the bloodstream as a marker of myocyte injury and necrosis(1–3). Their clinical presentations overlap substantially, with shared features including chest pain, electrocardiographic abnormalities, and regional wall motion abnormalities. However, their underlying pathophysiology differs markedly. AMI most commonly results from epicardial coronary artery occlusion due to atherosclerotic plaque rupture, whereas acute myocarditis represents an inflammatory process with diverse aetiologies, including viral infection, toxins, cancer therapies, and autoimmune, infiltrative, or genetic disorders(4, 5).

Cardiovascular magnetic resonance (CMR) is the reference standard for myocardial tissue characterisation owing to its high spatial resolution and multiparametric capabilities, including T2-weighted imaging for oedema, first-pass perfusion for quantifying myocardial blood flow and microvascular obstruction, parametric mapping for diffuse and focal myocardial fibrosis and/or inflammation, and late gadolinium enhancement (LGE) imaging to quantify focal myocardial injury and/or scar(6). The patterns of LGE on CMR differ between AMI and acute myocarditis. AMI typically demonstrates a subendocardial to transmural pattern in a culprit coronary artery distribution, whereas acute myocarditis more often shows a patchy subepicardial or mid-wall enhancement involving the lateral wall and septum(7, 8).

Traditionally, the extent of myocardial injury in both AMI and acute myocarditis has been inferred from the peak elevation of cardiac troponin, which is widely used as a surrogate marker of myocardial infarct size and injury burden. In AMI, higher peak troponin levels are associated with larger infarct size, worse left ventricular ejection fraction, and poorer clinical outcomes(9–11).

Despite the common use of troponin and LGE to assess myocardial injury, the relationship between the two may differ substantially between AMI and acute myocarditis due to their distinct pathophysiology. In AMI, troponin release is largely driven by transmural necrosis and its correlation with LGE extent is relatively strong, particularly when peak troponin is measured after reperfusion and when microvascular obstruction is considered(12). In acute myocarditis, troponin elevation reflects patchy inflammatory injury that may not correspond directly to LGE burden, especially in the acute phase, where oedema can influence enhancement patterns and potentially overestimate the true LV scar(13). Degree of peak troponin elevation can also be affected by several factors, including the timing of blood sampling post-event, the specific assay used, and the presence of comorbidities such as renal failure(14). To date, no studies have examined the correlation between peak troponin and LGE extent in both AMI and acute myocarditis using contemporary high-sensitivity troponin assays and standardised CMR protocols.

### Hypothesis

We hypothesised that the relationship between peak high-sensitivity troponin and the extent of myocardial injury measured by LGE on CMR differs between AMI and acute myocarditis. Specifically, we hypothesised that peak troponin would demonstrate a strong, linear correlation with LGE extent in AMI, reflecting irreversible myocyte necrosis, but a more variable relationship in acute myocarditis, where troponin release reflects patchy inflammatory injury and myocardial oedema that may not correspond directly to LGE extent.

### Aim

To quantify and compare the strength of association between peak high-sensitivity troponin and CMR-derived LGE extent in patients with AMI and those with acute myocarditis.

## METHODS

### Study Design

All patients aged ≥18 years with a confirmed diagnosis of AMI or acute myocarditis from April 2013 to April 2024 who underwent a CMR scan during their admission at two institutions (Royal North Shore Hospital, Sydney, Australia or North Shore Private Hospital, Sydney, Australia) were retrospectively recruited from the electronic health record. Baseline demographic, laboratory and CMR imaging parameters were collected. High-sensitivity cardiac troponin I was measured in all patients (Abbott Diagnostics, Abbott Park, IL, USA). The study was approved by the Northern Sydney Local Health District Human Research Ethics Committee and a waiver of individual written informed consent was provided due to low-risk retrospective nature of the study.

### Imaging Protocol

All studies were performed on 1.5 T and 3 T scanners from two vendors (Aera, Siemens Healthineers, Erlangen, Germany; Ingenia, Philips Healthcare, Best, The Netherlands, respectively) using standardised protocols(15). Cine images were acquired in the 2-chamber, 3-chamber, 4-chamber, and a contiguous stack of short-axis views using balanced steady-state free-precession (bSSFP) sequences. Phase-sensitive inversion-recovery (PSIR) and magnitude reconstruction LGE images were acquired ~10 minutes after intravenous administration of a gadolinium-based contrast agent (0.1–0.2 mmol/kg). Imaging was performed in contiguous short-axis slices covering the entire left ventricle and in standard long-axis views (2-, 3-, and 4-chamber). The inversion time was individually adjusted using a TI-scout sequence to null normal myocardium.

### Image analysis

CMR post-processing and analysis was performed off-line using 1CMR Pro (MyCardium AI Limited, London, UK). LV/RV volumes, mass and ejection fraction were quantified from short-axis cine images with semi-automated endocardial and epicardial smooth contour tracing. Papillary muscles were included in the LV cavity for volumetric measurements and excluded from myocardial mass. LGE extent was quantified using Segment (Version 4.1.0.1, Medviso AB, Lund, Sweden), using the expectation-maximization weighted intensity (EWA) algorithm in this software and shown in Figure 1. This EWA algorithm provides semi-automated delineation of hyperenhanced myocardium, as previously described(16). Regions of microvascular obstruction (MVO) were included in LGE extent.

**Figure 1.**
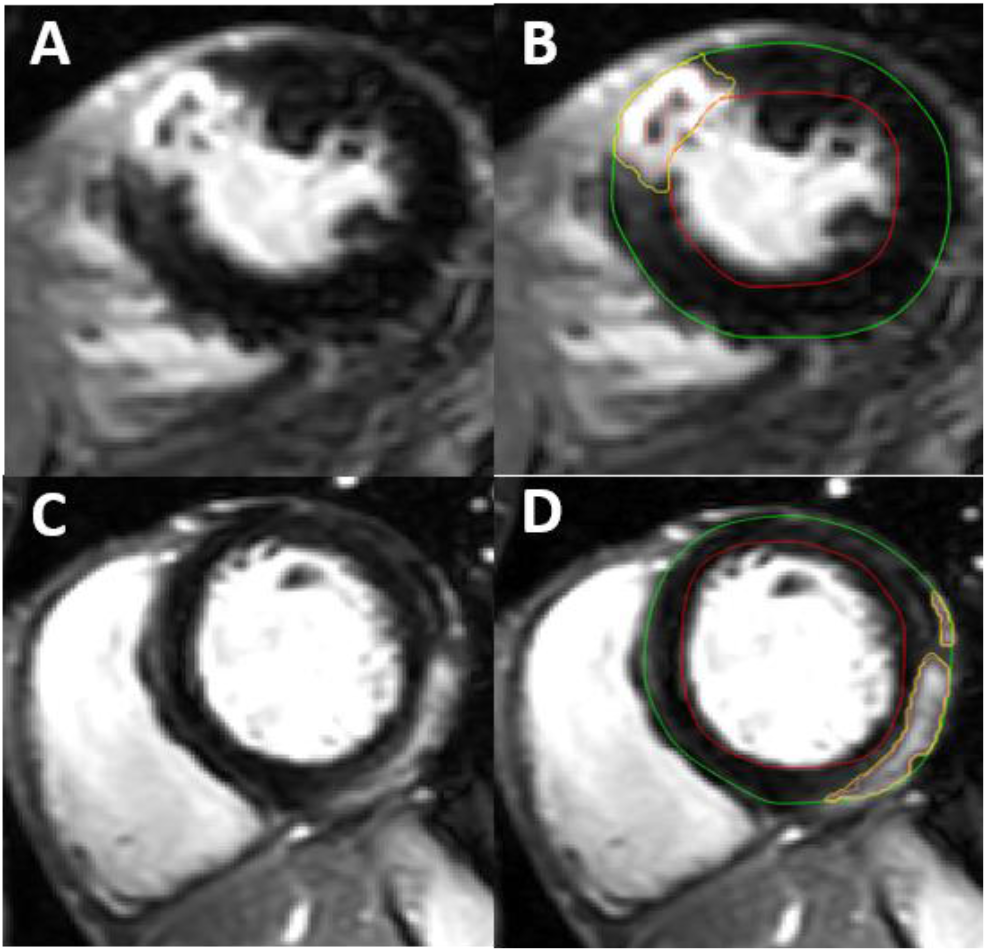
TOP ROW: A: Image showing transmural late gadolinium enhancement (LGE) and microvascular obstruction (MVO) in the anterior wall consistent with acute myocardial infarction (AMI), and B: typical segmentation by the EWA algorithm in the same patient BOTTOM ROW: C: Image showing subepicardial late gadolinium enhancement (LGE) in the inferolateral wall consistent with acute myocarditis, and D: typical segmentation by the EWA algorithm in the same patient

Automatic EWA segmentation of the hyper-enhancing region is delineated in yellow. Endocardium is delineated in red and epicardium in green.

### Statistical analysis

Baseline characteristics, laboratory data and CMR parameters were summarised with descriptive statistics. Continuous variables were presented as mean (standard deviation, SD) or median [interquartile range, IQR] and compared by student’s t test or Mann-Whitney U test depending on normality of distribution. Categorical variables were reported as frequencies (percentage) and compared by Fisher’s exact test. The relationship between changes in measured variables was examined by Pearson (r) or Spearman (rho) correlation based on normality of distribution. A p-value <0.05 was considered statistically significant. All statistical analyses were performed using SPSS Statistical software (IBM Corp. Released 2024. IBM SPSS Statistics for Windows, Version 30.0. IBM Corp, Armonk, New York).

## RESULTS

### Study Population and Demographics

Patient demographics, troponin and CMR parameters are listed in Table 1. The study cohort included 96 patients, divided into two groups: AMI (n=49) and acute myocarditis (n=47). The AMI group were older than those in the acute myocarditis group (p <0.001). Both groups were predominantly males (p=0.58) with no difference in BMI (p=0.42).

**Table 1.** Patient demographics, troponin and CMR parameters.

|  | Total (n=96) | AMI (n=49) | Acute myocarditis (n=47) | p-value |
| --- | --- | --- | --- | --- |
| **Demographics** |  |  |  |  |
| Male sex, n (%) | 69 (72) | 34 (69) | 35 (74) | 0.58 |
| Age, years | 51.9±19.3 | 62.0±13.7 | 41.3±18.7 | <0.001 |
| BMI, kg/m<sup>2</sup> | 25.8±4.4 | 26.3±4.7 | 25.3±4.0 | 0.42 |
| Height, m | 1.7±0.9 | 1.7±0.9 | 1.8±0.9 | 0.16 |
| Weight, kg | 78.8±17.0 | 78.9±16.0 | 78.7±18.2 | 0.80 |
| BSA, m2 | 1.9±0.2 | 1.9±0.2 | 2.0±0.3 | 0.82 |
| **Troponin** |  |  |  |  |
| Peak high-sensitivity troponin I, ng/L | 14493 [2324-39842] | 32470 [3109-104699] | 7295 [1857-22550] | 0.002 |
| **CMR parameters** |  |  |  |  |
| Time from peak troponin to CMR, days | 3 [2-6] | 4 [3-6] | 2 [1-4] | 0.001 |
| Time from presentation to CMR, days | 4 [2-6] | 5 [3-7] | 3 [2-6] | 0.01 |
| LVEDV, mL | 175 [149-205] | 180 [158-217] | 161 [142-192] | 0.04 |
| LVEDVi, mL/m2 | 92 [80-107] | 96 [82-113] | 88 [80-97] | 0.01 |
| LVESV, mL | 86 [67-108] | 102 [80-138] | 77 [62-95] | <0.001 |
| LVESVi, mL/m2 | 44 [37-58] | 55 [41-63] | 39 [35-47] | <0.001 |
| LVEF, % | 49 [42-56] | 45 [36-52] | 55 [49-58] | <0.001 |
| LVM, g | 119 [100-139] | 122 [102-147] | 114 [94-134] | 0.05 |
| LVMi, g/m2 | 60 [53-69] | 65 [55-74] | 57 [50-62] | 0.01 |
| RVEDV, mL | 156 [128-189] | 147 [124-176] | 164 [132-194] | 0.23 |
| RVEDVi, mL/m2 | 81 [67-95] | 78 [62-95] | 86 [72-96] | 0.11 |
| RVESV, mL | 71 [56-98] | 66 [55-97] | 79 [58-98] | 0.29 |
| RVESVi, mL/m2 | 36 [29-48] | 32 [28-48] | 40 [33-48] | 0.15 |
| RVEF, % | 54 [47-59] | 55 [43-60] | 53 [48-59] | 0.83 |
| LA area, cm<sup>2</sup> | 25 [21-28] | 25 [21-28] | 24 [20-27] | 0.27 |
| RA area, cm<sup>2</sup> | 23 [19-27] | 22 [19-26] | 23 [19-29] | 0.26 |
| **LGE details** |  |  |  |  |
| LGE extent, g | 15.5 [6.8-35.8] | 25.0 [13.0-56.0] | 10.0 [5.5-16.5] | <0.001 |
| LGE extent of total LVM, % | 13.5 [6.4-27.6] | 23.2 [9.6-41.7] | 10.1 [4.8-15.6] | <0.001 |
| Peak Troponin/LGE (g) ratio, ng/L per g | 1015 [315-2481] | 1299 [419-3233] | 909 [310-1446] | 0.02 |

Data presented as frequencies (%), mean±SD, or median [interquartile range].
Abbreviations: CMR, cardiovascular magnetic resonance; AMI, acute myocardial infarction; BMI, body mass index; BSA, body surface area; LVEDV, left ventricular end-diastolic volume; LVEDVi, indexed left ventricular end-diastolic volume; LVESV, left ventricular end-systolic volume; LVESVi, indexed left ventricular end-systolic volume; LVEF, left ventricular ejection fraction; LVM, left ventricular mass; LVMi, indexed left ventricular mass; RVEDV, right ventricular end-diastolic volume; RVEDVi, indexed right ventricular end-diastolic volume; RVESV, right ventricular end-systolic volume; RVESVi, indexed right ventricular end-systolic volume; RVEF, right ventricular ejection fraction; LA, left atrium; RA, right atrium; LGE, late gadolinium enhancement

### Cardiac Troponin

Peak high-sensitivity troponin I level was higher in the AMI group (p=0.002).

### CMR parameters

The interval from clinical presentation to CMR was longer in the AMI group (p = 0.01), as was the time from peak troponin to CMR (p = 0.001). Patients with AMI exhibited greater indexed end-diastolic left ventricular volumes (p = 0.01) and indexed left ventricular mass (p = 0.01), compared to those with acute myocarditis. LV ejection fraction was lower in the AMI group compared to the acute myocarditis group (p < 0.001). No differences between groups were observed in right ventricular end-diastolic and end-systolic volumes, RV ejection fraction and left atrial and right atrial areas.

### Tissue Characterisation

The extent of LGE, quantified in grams, was substantially greater in the AMI group (p <0.001). LGE, as a percentage of total LV mass, was also higher in the AMI group (p < 0.001). Additionally, the ratio of peak troponin to LGE mass was significantly higher in the AMI group (p = 0.02), as shown in Figure 2. There was a moderate positive correlation between peak troponin levels and LGE extent when analysed by subgroup: AMI Spearman’s rho = 0.56, p < 0.001, and acute myocarditis Spearman’s rho = 0.58, p <0.001. The correlation matrix of peak troponin to LGE extent in AMI and acute myocarditis is also shown in Figure 2.

**Figure 2.**
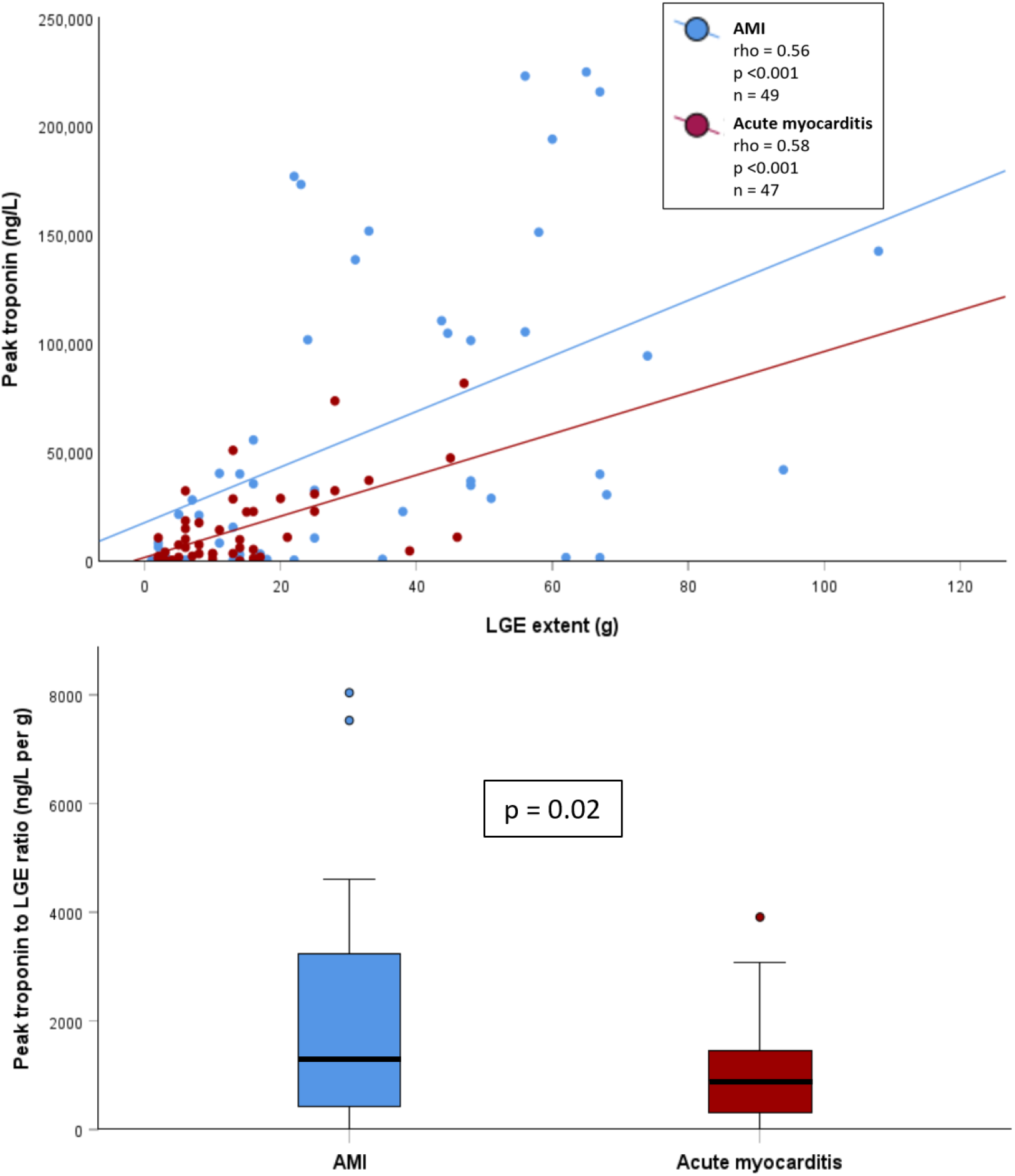
TOP PANEL: Spearman (rho) correlation matrix of peak troponin and late gadolinium enhancement (LGE) extent in acute myocardial infarction (AMI) and acute myocarditis BOTTOM PANEL: Distribution of peak troponin to late gadolinium enhancement (LGE) ratio (ng/L per g) in acute myocardial infarction (AMI) and acute myocarditis

## DISCUSSION

This study evaluated the relationship between peak cardiac troponin and the degree of myocardial injury as defined by CMR-LGE extent in patients with AMI and acute myocarditis. The principal finding is that AMI typically has an approximately 40% greater amount of troponin release per unit LGE mass compared to acute myocarditis. Also, peak troponin levels correlated positively with LGE extent in both conditions, but the magnitude of LGE and LV systolic dysfunction was greater in AMI.

Our study reinforces the strong association between cardiac troponin release and infarct size in AMI that has been demonstrated previously(11, 17). In our cohort, AMI patients had markedly higher peak troponin, larger LGE mass, and more adverse LV remodelling with higher indexed LVED volumes and lower LVEF than patients with acute myocarditis. In CMR studies, troponin has correlated closely with LGE-defined infarct size and microvascular obstruction (MVO). Though not specifically quantified in our study, MVO is frequently observed in larger infarcts, can further increase troponin and is associated with poorer outcomes(18).

In acute myocarditis, the relationship between troponin and LGE is more heterogeneous due to patchy or diffuse distribution of inflammation, rather than focal necrosis and fibrosis alone(19). In their cohort of patients with acute myocarditis and preserved ejection fraction, Acquaro et al. found that an antero-septal pattern of LGE was associated with a high troponin release and worse prognosis(7). Our study found a moderate correlation between peak troponin and LGE extent in acute myocarditis, supporting troponin as a surrogate marker for global inflammatory burden(20). Despite this, patients with acute myocarditis had smaller LGE masses, relatively preserved LV systolic function, and lower LV volumes than patient with AMI. The predominantly subepicardial or mid-wall LGE distribution and the reversible component of oedema in acute myocarditis likely contribute to preserved LV function despite marked troponin elevation(5, 7, 19, 21).

An important aspect of this work is the evaluation of the ratio of peak troponin to LGE mass. The higher troponin-to-LGE ratio in AMI suggests greater circulating troponin per gram of enhanced myocardial tissue. This may reflect necrosis of dense compacted myocardium in AMI, whereas a combination of necrosis, interstitial expansion, and oedema in acute myocarditis(13). Re-perfused AMI also often shows a rapid, high troponin peak, while acute myocarditis may follow a more prolonged release profile(2, 22). Our findings extend this concept by demonstrating that a simple troponin-to-LGE mass ratio differs markedly between AMI and myocarditis. Taken together, these mechanisms provide a biological explanation for the distinct troponin–LGE relationships observed in the two conditions.

Clinically, these findings have important implications for the interpretation of troponin in acute myocardial injury. For a given troponin range, patients with AMI had larger LGE masses, higher LV volumes, and lower LVEF than patients with acute myocarditis, consistent with the heavier functional burden of transmural ischaemic scar compared with non-ischaemic scar. Prior CMR studies in ischaemic and non-ischaemic cardiomyopathies have shown that LGE extent and distribution are associated with adverse remodelling, impaired LV function, and higher risk of major adverse cardiovascular events(23–25). In acute myocarditis, subepicardial LGE and oedema often regress with improvement of wall motion abnormalities and generally favourable outcomes. Persistent LGE, however, may confer higher long-term risk of arrhythmia and heart failure(19). By contrast, ischaemic LGE usually represents permanent scar underlying chronic LV dysfunction, heart failure progression, and arrhythmic substrate. We observed that AMI is characterised by both larger LGE burden and more pronounced LV dysfunction than acute myocarditis for similar troponin ranges – reinforcing the prognostic value of integrating biomarker data with CMR myocardial tissue characterisation.

This study also underscores the importance of CMR timing when assessing troponin–LGE association. Oedema and LGE evolve dynamically in the days to weeks post-event and prior work suggests that the correlation between troponin and LGE in acute myocarditis is strongest when CMR is performed within approximately 5 days of troponin peak(26). In our cohort, patients with AMI underwent CMR typically two days later than patients with acute myocarditis, which may have attenuated the oedema-related LGE in the former.

Larger prospective studies with standardised CMR timing, LGE quantification methods and assessment for major adverse cardiac events are needed to validate the diagnostic and prognostic utility of the troponin-LGE ratio and to define practical cut-off values.

## Limitations

There are several limitations to our study. The retrospective nature of the study with a relatively small sample size (n=96) limits the generalisability. The timing of CMR differed between the two groups, with patients with AMI scanned two days later than those with acute myocarditis. As myocardial oedema and LGE are dynamic processes that can evolve rapidly in the acute phase, this temporal difference could influence the quantification of myocardial injury. Also, LGE extent could be overestimated given the absence of a follow up scan at an appropriate time interval following resolution of oedema. The quantification of LGE is also dependent on method. Whilst we employed the Expectation-Maximization Weighted Intensity algorithm (EWA), other techniques such as full-width at half-maximum and n-standard deviation methods might yield different results(27, 28). Notably, EWA was used in this study since it has an advantage over other methods that quantify voxels in a binary fashion based on a threshold, as wholly infarcted or not. Importantly, EWA quantifies the fraction of a given voxel that is damaged by weighting the injured volume according to signal intensity. This has validated accuracy in AMI(16), but has not been systematically applied in acute myocarditis. Regardless of this shortcoming in validation, all patients were analysed with the same method, which strengthens the robustness of the analyses.

## CONCLUSION

Although peak troponin and LGE extent are moderately correlated in both AMI and acute myocarditis, the absolute burden of myocardial injury and the relationship between biomarker release and LGE mass differ substantially between these disease entities. These findings highlight important pathophysiological differences in myocardial injury and suggest that troponin-based estimates of injury size are not directly interchangeable between AMI and acute myocarditis.

## Data Availability

All data produced in the present study are available upon reasonable request to the authors

## Notes

### Competing Interest Statement

The authors have declared no competing interest.

### Funding Statement

This study did not receive any funding

### Author Declarations

The study was approved by the Northern Sydney Local Health District Human Research Ethics Committee and a waiver of individual written informed consent was provided due to low-risk retrospective nature of the study.

